# A model assessing potential benefits of isolation and mass testing on COVID-19: the case of Nigeria

**DOI:** 10.1101/2020.09.01.20186288

**Authors:** Faraimunashe Chirove, Chinwendu E. Madubueze, Zviiteyi Chazuka, Sunday Madubueze

**Affiliations:** University of Johannesburg, Department of Mathematics and Applied Mathematics, South Africa; Federal University of Agriculture, Department of Mathematics, Statistics and Computer Science, Nigeria; Chinhoyi University of Technology, School of Natural Sciences and Mathematics, Zimbabwe; Dalhatu Araf Specialist Hospital, Department of Family Medicine, Lafia, Nigeria

**Keywords:** COVID-19, Isolation, Mass testing

## Abstract

We consider a model with mass testing and isolation mimicking the current policies implemented in Nigeria and use the Nigerian daily cumulative cases to calibrate the model to obtain the optimal mass testing and isolation levels. Mathematical analysis was done and important thresholds such the peak size relation and final size relation were obtained. Global stability analysis of the disease-free equilibrium indicated that COVID-19 can be eradicated provided that 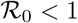 and unstable otherwise. Results from simulations revealed that an increase in mass testing and reduction of transmission from isolated individuals are associated with benefits of increasing detected cases, lowering peaks of symptomatic cases, increase in self-isolating cases, decrease in cumulative deaths and decrease in admissions into monitored isolation facilities in the case of Nigeria.

## 1 Introduction

The novel coronavirus disease 2019 (COVID-2019) is the world’s recent pandemic. It is a highly contagious disease caused by severe acute respiratory syndrome coronavirus-2 (SARS-CoV-2) [18,31] that belongs to the same family with severe acute respiratory syndrome coronavirus (SARS-CoV), and Middle East respiratory syndrome coronavirus (MERS-CoV) which are known to cause pneumonia in humans [18,22,2,33]. The first outbreak in Wuhan, China in December 2019 has since spread to the rest of the world [2,33,26] which saw it being declared a world pandemic in 2020 [22,27]

Currently, there are over 21 million confirmed cases, over 750 thousand deaths and close to 14 million recoveries. Africa alone has over one million confirmed cases, about 24 thousand deaths and about 780 thousand recoveries. Nigeria currently is the third highest contributor to the African burden with about 48 thousand confirmed cases, close to a thousand deaths and over 34 thousand recoveries [35]. The COVID-19 primary source of transmission was firstly suspected to be from wild animals to humans but in Nigeria and the rest of Africa, the primary source was patients infected with COVID-19 travelling into the continent [18,2,31]

In general, COVID-19 case fatality is low but people with co-morbidities like diabetes mellitus, cancer, chronic obstructive pulmonary diseases, and hypertension are at high risk of COVID-19 fatalities. The routes of transmission for SARS-CoV-2 are respiratory droplets (generated via sneezing and coughing), close contact with an infected person, contact with environmental surfaces and aerosols. Successful infection by a virus occurs when adequate viral load enters the body, penetrates the target cells, replicates and gets released to infect more cells [18]. The symptoms of COVID-19 illness are fever, dry cough, and fatigue, headache, haemoptysis, diarrhoea, difficulty with breathing and muscular pain [2,26,25]. Severe disease is characterised by acute respiratory syndrome, acute cardiac failure, acute kidney failure, anaemia, stroke, multiple organ dysfunction, and secondary bacterial infection [36]. The current preventive methods, among others, are maintaining physical distancing, observing cough etiquette, quarantine, isolation and community lock downs [33,34].

The incubation period of SARS-CoV-2 has been estimated to range from 2 to 14 days [22]. Available evidence showed that COVID-19 human-to-human transmission occurs during the asymptomatic incubation period which is usually 2 to 10 days [27]. This suggests that for any systems to control the infection effectively, the asymptomatic individuals should be subjected to testing so that preventive measures such as self and mandatory isolation can be effected to reduce the spread of the infection. Community mass testing is one of the strategies that can be used to identify infected individuals. Isolation is the separation of persons with contagious disease from those not infected [7]. The infected individuals can be isolated in hospital settings for severe and critical diseases while sub-clinical infections can stay at home. For isolation to be effective in halting transmission, cases should be identified before the onset of viral shedding or at least before the onset of peak viral shedding [33,14]. One of the effective testing instruments is the use of real time polymerase chain reaction (RT-PCR) [18,3] which was used effectively among sheltered people in Boston, USA and South Korea on the asymptomatic population to reduce the risk of community transmissions instead of focusing only on those with severe symptoms or high risk groups [3]. It has been argued that a population-wide mass testing would help identify individuals that are likely to transmit disease long before symptoms appear [28]. Thus, the fight against COVID-19 is a war against an invisible enemy which requires rapid detection of those that can transmit the infection and quickly isolate them before viral shedding commences.

COVID-19 sufferers can be classified into three groups: sub-clinical (asymptomatic and mild symptomatic) infections, severe disease, and critical disease [22]. The first two groups accounts for approximately 81% of cases and usually resolve without any pharmacologic intervention[22]. They pose serious danger to containment because they can transmit the virus without being aware of their status. This group often refuses isolation from the community and even when isolated, may attempt to escape from the isolation centre and can recover without any intervention. Those with severe disease account for 14% and would require hospitalization, supportive and pharmacologic therapies while those with critical disease account for the remaining 5% requiring nursing in the intensive care unit with ventilator [22]. Treatment administration remains largely supportive therapy [22,10].

Our study seeks to assess the potential impact of isolation and mass testing of the population on the transmission of COVID-19 in Nigeria using mathematical modelling approach. We present the background setting for Nigeria henceforth.

### 1.1 The Nigeria situation

Nigeria has a population of 205, 416, 152 persons as of 14 May 2020 [35]. The Nigeria COVID-19 cases started with an imported case of a 44-year-old Italian citizen who presented himself to the staff clinic on the 26*^th^* of February 2020 and was later confirmed as the first official case of COVID-19 in Nigeria on the 27*^th^* of February 2020. Contact tracing for the index case identified about 40 asymptomatic who were tested and produced one confirmed positive making it the second case of COVID-19 on the 8*^th^* of March 2020. More imported cases were reported in March and since then the infection rose to current aforementioned pandemic levels in Nigeria [21,12].

The first COVID-19 related death was recorded on the 22*^nd^* of March 2020 and the fatality was due to co-morbidities complications. Cumulative confirmed cases in Figure 1 are used for calibration of the mathematical model. The lock down was announced by Nigeria Government effected measures such as travel bans from his risk countries and as well as lock down in order to halt the community spread of the COVID-19. The initial lock down was in Lagos, Abuja, and Ogun states on the 30*^th^* of March 2020 and later extended to other states. Subsequent easing of lock down measures ushered in new measures including mandatory wearing of face masks, nationwide night-time curfew and a ban on non-essential travelling between different states of the country being enforced [1]. Mass testing is yet to take effect in Nigeria. As of 22 August 2020, Nigeria has carried out 374,077 sample tests which is small for a country with over 200 million people [11]. This poor testing capacity makes the epidemic curve not flatten in Nigeria. Nigeria has about 61 laboratories for COVID19 testing. The testing is for three categories of people, those with travelling history outside Nigeria and symptoms within 14 days of arrival, those with contact with confirmed case and symptoms, and those with symptoms and stay in high prevalence of COVID-19 area in Nigeria. [11,30]

**Fig. 1.**
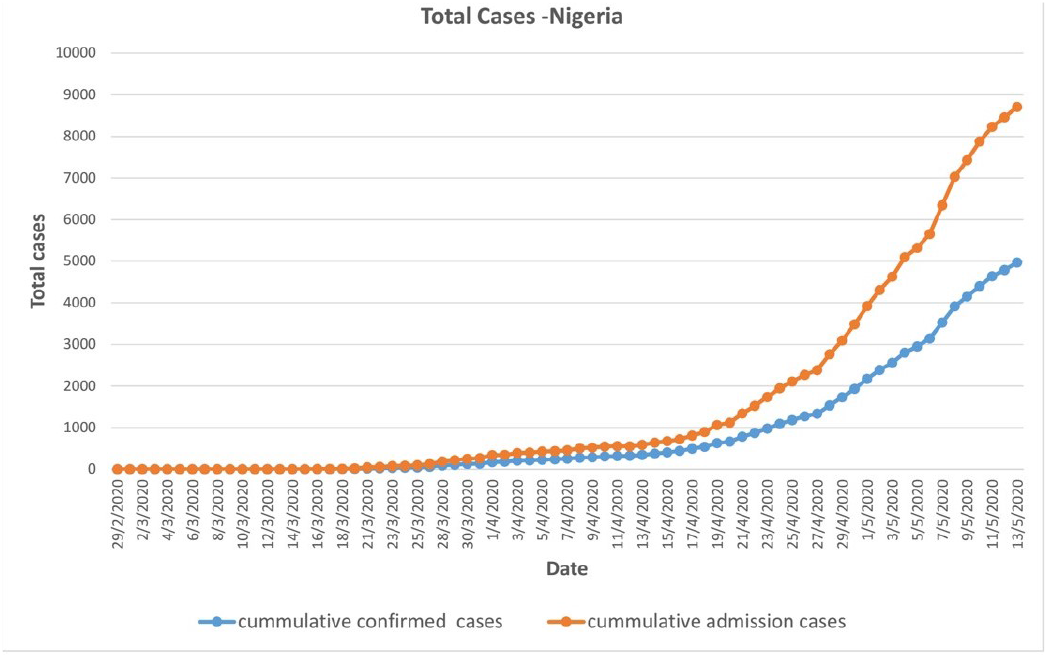
Cumulative cases in Nigeria

### 1.2 Mathematical modelling of COVID-19

A number of mathematical models for COVID-19 have so far been established which looked at the different effects of intervention strategies and predict the probable outcomes of the pandemic. The Studies presented in [6,20,15,32,9,19,16,24,23,8] used the standard SEIR compartmental structure to model the various aspects of COVID-19 dynamics interventions. However, the studies in [5,17] points out the inadequacy of the framework and presented improved frameworks which included the quarantined classes as well as the dead compartments. The results from all the aforementioned studies focused on the benefits of control measures such as quarantining, travel restrictions and social distancing, among others but did not take into consideration the effects of the asymptomatic infectious individuals as these are one of the major undetected cases that are drivers of infection. To detect this group of individuals, measures such as mass testing and isolation will need to be implemented. Checchi *et al*. [8] presented a discrete time age structured mathematical model for the spread of COVID-19 in African settings looking at the effects of isolation, social distancing and shielding in the reduction of COVID-19 cases in Africa using data from Mauritius, Nigeria and Niger. They concluded that isolation and social distancing were effective when subjected to lock down conditions. Our current work on the dynamics of the novel COVID-19 virus in Nigeria uses a deterministic modelling approach to estimate the potential impact of self and mandatory isolation, and mass testing. The organisation of the paper is as follows: in section 2 we present the model formulation of our deterministic mathematical model for COVID-19 dynamics in Nigeria, section 3 presents fundamental properties of the model, threshold computation and equilibrium stability analysis.

## 2 Materials and Methods

We formulate a mathematical model to study the dynamics COVID-19 incorporating mass testing and, self and mandatory isolation. We consider a homogeneously mixing population that comprises of the following compartments: the susceptible population *S*(*t*), the exposed population with the virus but not yet able to transmit it to other individuals *E*(*t*), infectious asymptomatic population not yet tested *I_A_*(*t*), infectious asymptotic population that has been tested and self isolating *I_AT_*(*t*), infectious symptomatic population *I_s_*(*t*), infectious symptomatic isolated population under health care enclosures and monitoring *J*(*t*) and the removed population *R*(*t*). We assume that mass testing mostly affects those in classes *I_A_*(*t*), *I_s_*(*t*) mainly because these are the only classes where individuals test positive. Individuals in *S*(*t*) and *E*(*t*) and *R*(*t*) will test negative and thus, will remain in the same class as the ones untested. Individuals in *I_s_*(*t*) as soon as they test positive are immediately transferred into isolation centres and hence, are assumed not to self isolate. The total human population *N*(*t*) is

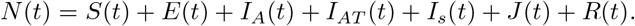

Since COVID-19 is a respiratory, we leave out the demographic parameters and concentrate on the infection and intervention dynamics due to differences in time scales of occurrence between demographic and infection dynamics. The force of infection for the model is assumed to be

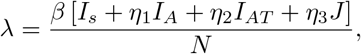

where *β* is the effective transmission rate of COVID-19 and *η*_1_, *η*_2_, *η*_3_ are modification parameters where 1 >*η*_1_ >*η*_2_ >*η*_3_ ≥ 0 which quantify the contribution of each infectious class compared to the most infectious ones. Thus, individuals that are symptomatic and not tested or isolated in class *I_s_* are more infectious than individuals in class *I_AT_* and those in class *J*. Exposed individuals *E*(*t*) will progress into the *I_A_*(*t*) and *I_s_*(*t*) at rates *σ*(1 − *ϵ*) and *σϵ* respectively where *σ* is the rate of progression and *ϵ* ∈ [0, 1] is the proportion of individuals from *E*(*t*) that develop symptoms. Due to mass testing, some individuals in *I_A_*(*t*) test positive and are required to self isolate progressing to the class *I_AT_* (*t*) with a probability rate of detection due to mass testing *γ* and they can recover from the infection and move to the removed class *R* at a rate *ω*. Those in class *I_s_*(*t*) of symptomatic individuals either progress to mandatory isolation at a rate *δ* = *γ* + *ϕ* or recover from the infection at rate *ζ*. Here, *ϕ* is the isolation rate due to the individuals visiting the health care centres before mass testing detection and *γ* is the probability rate of detection due to mass testing of *I_s_* population. They can also die due to COVID-19 induced death at a rate *d*_1_. Individuals in the *I_AT_* (*t*) class can either progress to the mandatory isolation care centers after developing symptoms at a rate *pδ*, where *p* is the probability that an individual in *I_AT_* (*t*) develops symptoms. They can also recover at a rate *α* and move to *R*. Individuals in the *J* class can either recover at a rate *ν* or die due to the virus at a rate *d*_2_. The flow diagram of the model with the aforementioned assumptions is in Figure 2 and the governing equations are given by systems of equations (1).

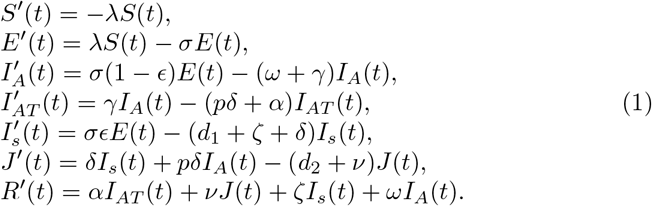

**Fig. 2.**
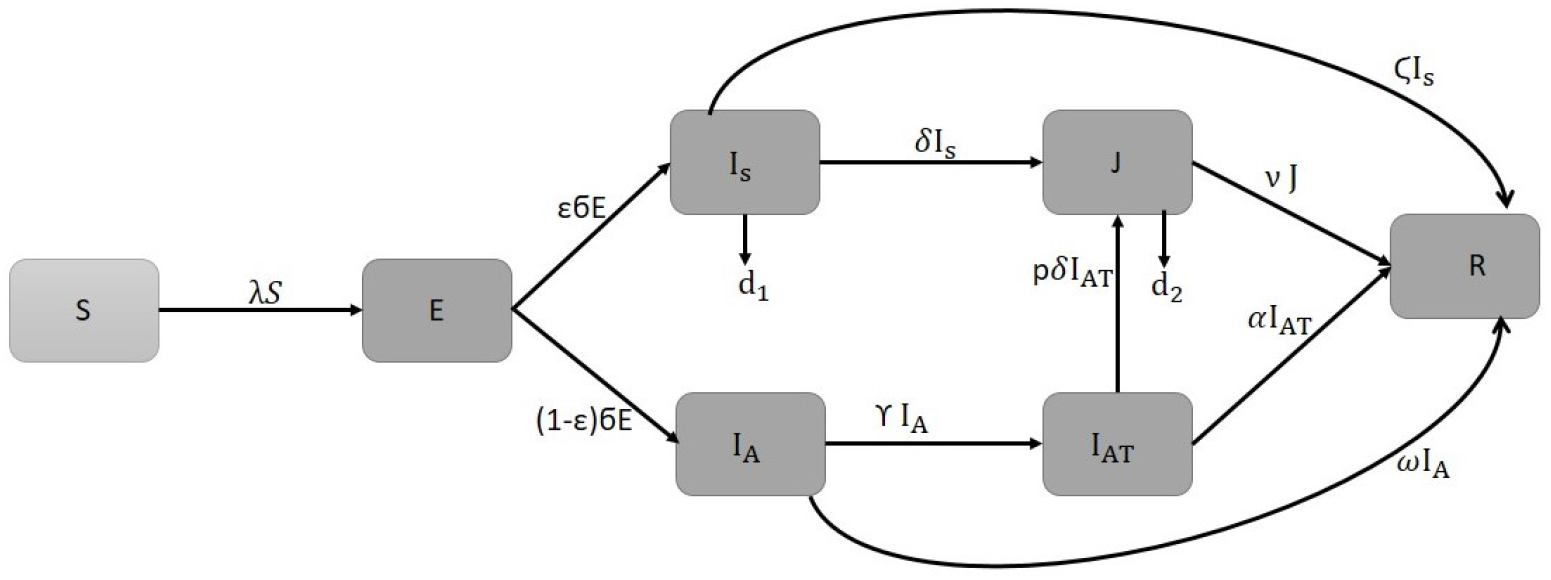
Model flow diagram for the dynamics of COVID-19

The model (1) has initial conditions given by

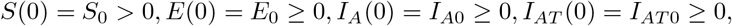

*I_s_*(0) = *I_s_*_0_ ≥ 0, *J*(0) = *J*_0_ ≥ 0, *R*(0) = *R*^0^ ≥ 0 and based on biological considerations, all feasible solutions of model (1) exist in the region

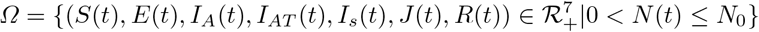

## 3 Results

### 3.1 Mathematical results

#### 3.1.1 Positivity and boundedness of solutions

Model (1) describes dynamics of COVID-19 within the human population and therefore it is important and necessary to prove that all the variables used in the model (*S*(*t*), *E*(*t*), *I_A_*(*t*), *I_AT_* (*t*), *I_s_*(*t*), *J*(*t*), *R*(*t*)) are non-negative for all time and also that solutions for model system (1) with positive initial data will remain positive for all time and are bounded in *Ω*.

##### Theorem 1

*The solutions (S(t), E(t), I_A_(t), AT(t), I_s_, J(t), R(t)) of model (1) with nonnegative intial conditions are positive for all t* ≥ 0.

###### Proof

Solving the first equation of model system (1) using the separation of variables method we have

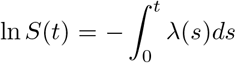

which simplifies to

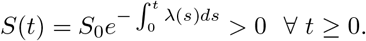

It is easy to follow a similar analysis to show that the other state variables *E*(*t*), *I_A_*(*t*), *I_AT_* (*t*), *I_s_*(*t*), *J*(*t*), *R*(*t*) are non-negative for all *t* > 0. Based on this we conclude that the solutions of model system (1) remain positive for all *t* ≥ 0 and this completes the proof.

##### Theorem 2

*All solutions* (*S*(*t*), *E*(*t*), *I_A_*(*t*), *I_AT_* (*t*), *I_s_*(*t*), *J*(*t*), 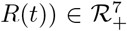 *of model system (1) are bounded in Ω*.

Proof

Considering the total population for model system (1) *N*(*t*) we have

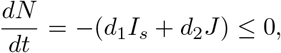

by the positivity of *I_s_* and *J*. The solution to the differential inequality using intial conditions *N*(0) = *N*_0_ is given by

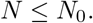

Hence, we conclude that *N*(*t*) is bounded above and all state variables are bounded.

The region *Ω*, in which the solutions of model system (1) are restricted is therefore a feasible region.

#### 3.1.2 Equilibrium analysis and the Reproduction number

The disease free equilibrium point is given by

*E*_0_ =(*N*_0_, 0, 0, 0, 0, 0, 0). Using the next generation matrix approach by [29], our model satisfy the conditions *A*1 to *A*5 and the basic reproduction number 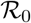 is given by

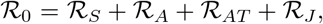

where

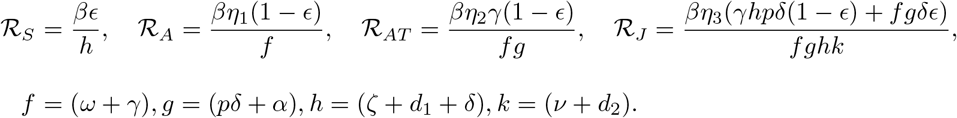

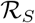 is a local reproduction number that measures the secondary infections that arise when the index case is a symptomatic infectious individual, 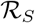 is a local reproduction number that measures the secondary infections that arise when the index case is an asymptomatic infectious individual not tested, 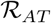 is a local reproduction number that measures the secondary infections that arise when the index case is an asymptomatic infectious individual who has been tested and self isolating and 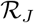, is a local reproduction number that measures the secondary infections that arise when the index case is an isolated infectious individual.

It follows from the fact that the model satisfy conditions *A*1 to *A*5 of [29] that the disease free equilibrium is stable when 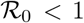 and unstable when 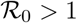. Hence, the stability theorem below holds

##### Theorem 3

*The disease free equilibrium point E_0_ for model system (1) is locally asymptotically stable provided that* 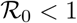.

#### 3.1.3 Final and peak size relation

In relation to controlling the novel COVID-19 virus, isolation has been one of the major worldwide control measure with most countries enforcing lock downs to the general susceptible population and mandatory isolation for infected individuals. In epidemic modelling quantities such as the reproduction numbers 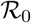, 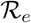, peak size and final peak size of the epidemic, are important in providing epidemiologists with useful information on the effects of such control measures on the dynamics of infection within the population [13]. We consider an epidemic that started as a result of an infected visitor from outside the population. The population is a completely susceptible population of size *N* ≈ *S*_0_. We use Feng *et al*. [13] to calculate the peak size of the pandemic and the simple Kermack-Mckendrick model approach in the work by Fred Brauer [4] to calculate the final size relation for COVID-19 as shown below. The population is a completely susceptible population of size *N* ≈ *S*_0_.

##### Peak size relation

To determine the peak of the epidemic we define a weighted infected sum *y*(*t*) as

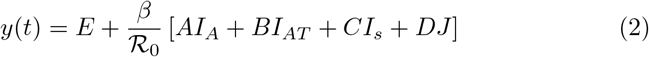

where

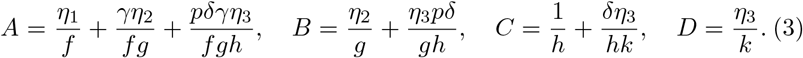

The infected compartments *E*(*t*), *I_A_*(*t*), *I_AT_* (*t*), *I_s_*(*t*) and *J*(*t*) are considered since they contribute to the infection. Now differentiating (2) with respect to *t* yields

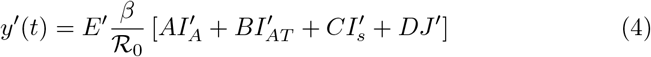

and substituting E′(*t*), 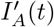, 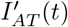, 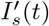 and *J*′(*t*) from system (1) into (4) and simplifying yields

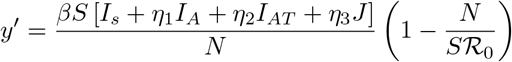

which is equivalent to

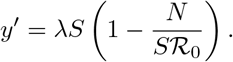

Hence, dividing *y*′(*t*) by *S*′(*t*) of model system (1) yields

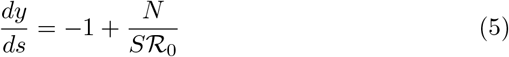

where *N* = *N*_0_ is a constant considering the limiting population of *N*. Integrating (5) with initial conditions *y*(0) = *y*_0_ and *S*(0) = *S*_0_ gives

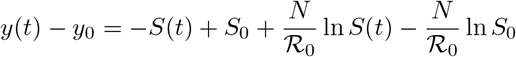

which simplifies to

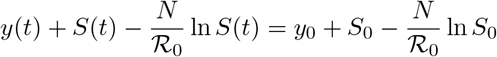

where 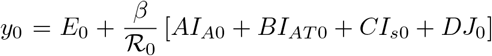. The maximum value of *y*(*t*) at any time *t* is the number of infective when *y*′(0) = 0, that is when 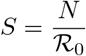. This is given by

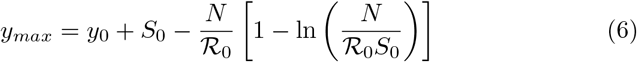

and this is obtained when 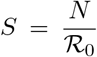 and 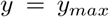 in equation (6) then it follows that equation (6) is the peak of the epidemic size.

Final size relation

To determine the final size relation of the basic reproduction number and the size of the epidemic, we take *S*_0_ = *N* and *S*_∞_ to be a non-negative smooth decreasing function that tends to a limit as *t* →∞, i.e *S*_∞_ > 0. Using the Kermack-Mckendrick model approach [4], let

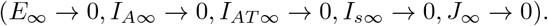

Adding the first two equations and, also the first three equations of system of equations (1), we obtain

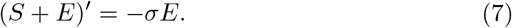

Integrating equation (7) on [0, ∞), we obtain

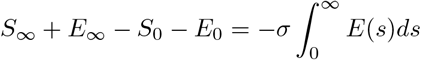

but *S*_0_ = N, *E*_∞_ = 0 hence we have

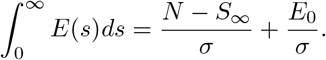

Integrating equation three of system equation(1) we have

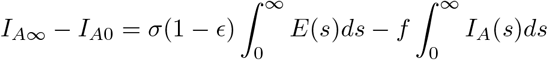

so

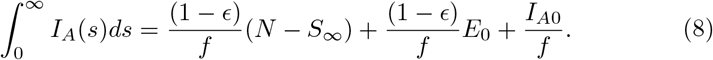

Similarly

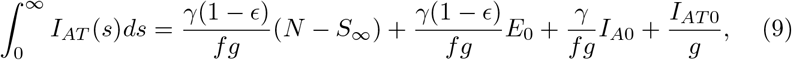

and

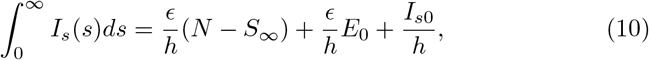

and also

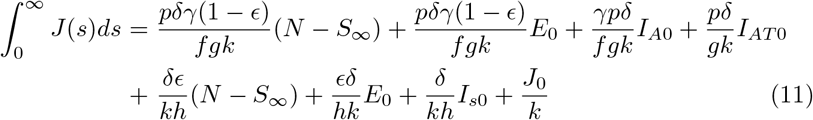

From the first equation of system (1) we have

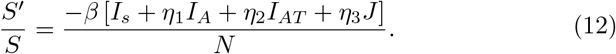

Therefore, integrating (12) on [0, ∞) yields

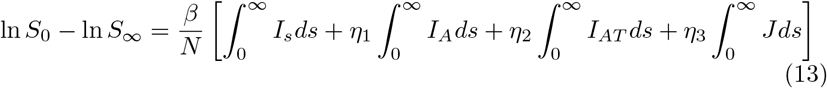

and hence substituting (8)-(11) into (13) and simplifying yields

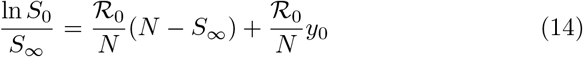

where 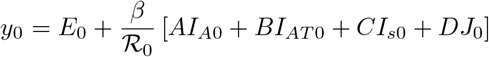. Therefore equation (14) gives the final size relation with initial terms *I_s_*_0_, *E*_0_, *I_A_*_0_, *I_AT_*_0_ and *J*_0_. If the initial terms are assumed to be *E*(0) = *I_A_*(0) = *I_AT_* (0) = *I_s_*(0) = *J*(0) = 0, and if a small number of infectives are introduced into the population then we have *S*_0_ ≈ *N*_0_ such that the final size relation has the form

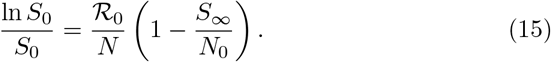

Equation (15) can be simplified further to give

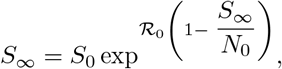

therefore 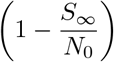 is the clinical attack rate and (*S*_0_ −*S*_∞_) is the size of the epidemic (that is the number of individuals in the population who are infected over the course of the epidemic).

## 4 Global stability analysis of the disease free equilibrium

### Theorem 4

*The disease free equilibrium for models system (1) is globally asymptotically stable provided that* 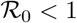.

#### Proof

In order to prove the above theorem we construct a Lyapunov function as follows

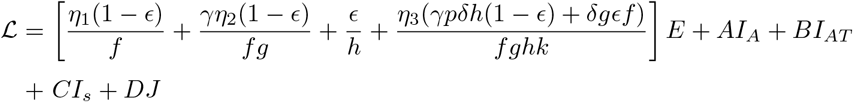

where *A, B, C* and *D* are defined in equation (3). Differentiating 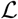 yields

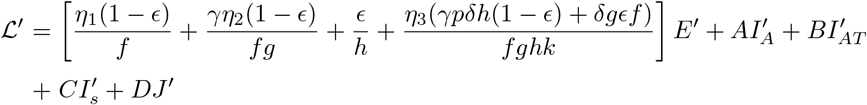

which simplifies to

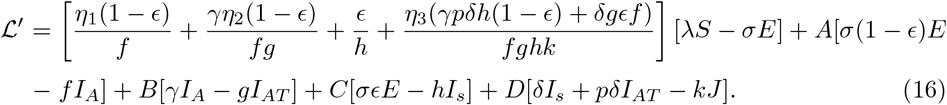

Therefore expanding (16) and simplifying with the definition of *A, B, C* and *D* in equation (3) yields

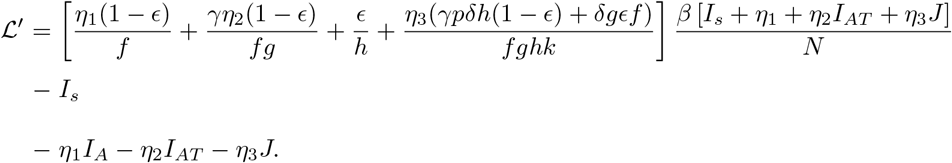

Using the definition of 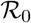 this further simplifies to

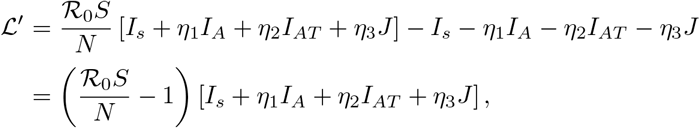

but 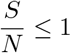 in the domain such that it follows that

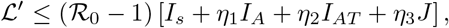

therefore is 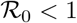, then it follows that 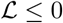. It also follows that when *E*(*t*)= *I_A_*(*t*)= *I_AT_* (*t*)= *I_s_*(*t*)= *J*(*t*)=0, 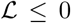 and hence the largest compact invariant set is the singleton {*E*_0_}. Therefore by LaSalle’s invariance principle it follows that *E*_0_ is globally asymptotically stable in *Ω*. This completes the proof.

### Numerical simulations results

We present simulations to assess the potential impact of isolation and mass testing on COVID-19 by fitting the model to cumulative cases in Nigeria. Data on cumulative cases was obtained from official public sources from the Nigeria government [11]. Our simulations will focus on two parameters *γ* which is the mass testing parameter and *η*_2_ the measures of the impact of contribution of the asymptomatic self-isolating population. The testing parameter *γ* assumes the values 0, 0.0261 and 0.0496. The value zero represents absence of mass testing after day 87, The value 0.0261 represents the baseline fitted value which we take as the base testing level by day 87 and the value 0.0496 represents an increase of testing level after day 87. The parameter *η*_2_ assumes the value 0.5391 as the baseline fitted value. *η*_2_ = 0 represents no transmission of COVID-19 by self-isolating individuals after day 87 and *η*_2_ =0.0539 represents reduced transmission rate of COVID-19 from the baseline contribution. Figures 7, 8 and 9 show the model projections for COVID-19 from day one of infection up to day 365. The pair (*γ*, *η*_2_) = (0, 0) is a scenario where the mass testing is stopped and no transmission from self-isolating individuals after day 87 and (*γ*, *η*_2_) = (0.0496, 0.0539) is a scenario with increased mass testing and reduced transmission from self-isolating individuals after day 87 whilst (*γ*, *η*_2_) = (0.0261, 0.5391) is the baseline pair from data fitted to the model.

Figure 3 shows the fit for cumulative cases for Nigeria from day one (1) of first infection recorded to day 87 of recorded infections. The model fit agrees relatively to the data and hence, we use the parameter values for this fit to make the projections on the impact of mass testing and isolation in Nigeria. The fitted parameter values are presented in Table 1.

**Fig. 3.**
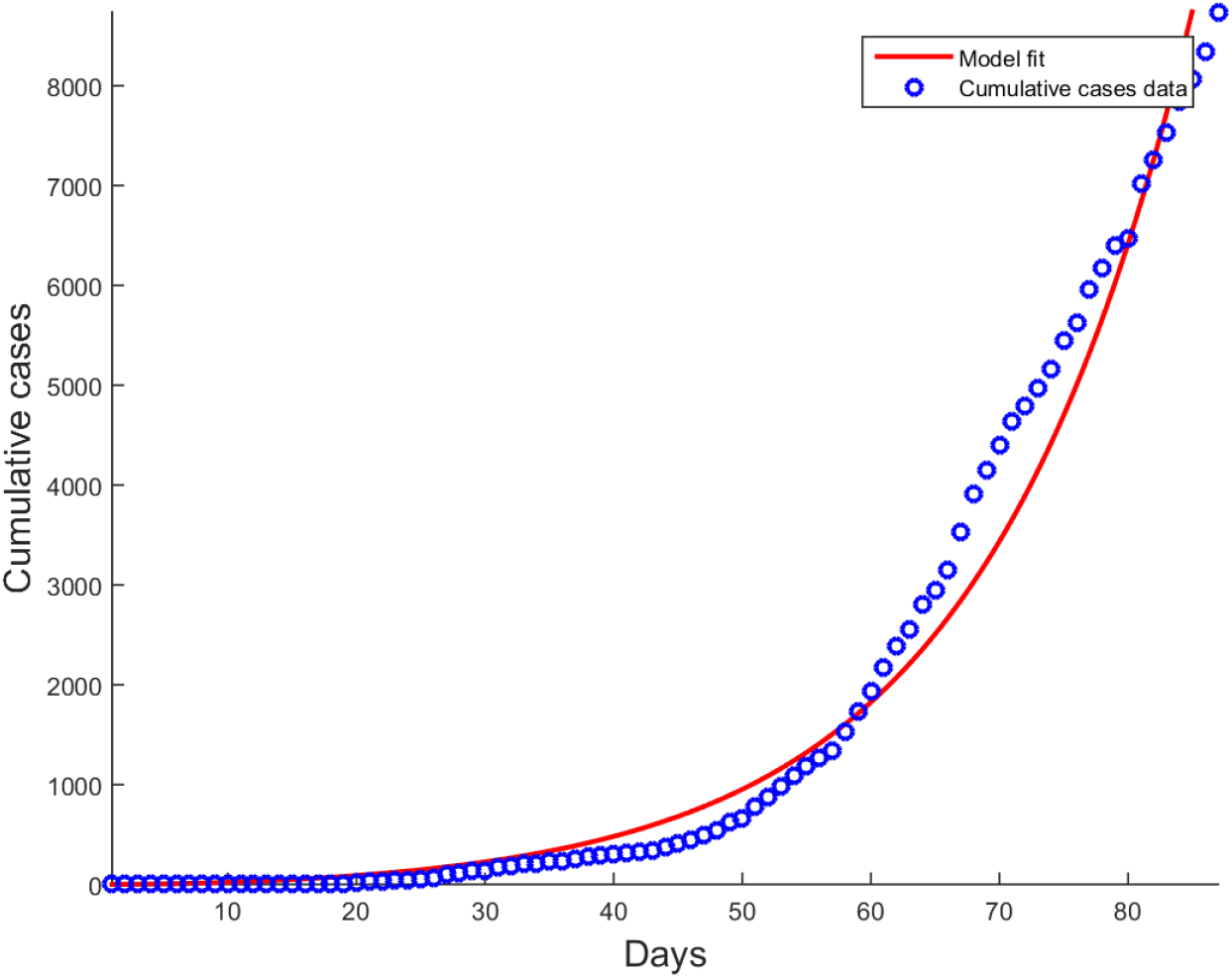
Model fit for COVID-19 Cumulative cases for Nigeria since day one of infection to day 87.

**Table 1.** Parameter values for the model with mass testing and isolation

| Parameter | Value | source |
| --- | --- | --- |
| $\beta$ | 0.9999 | fitted |
| $\eta_1, \eta_2, \eta_3$ | 0.9577, 0.5391, 0.9144 | fitted |
| $\sigma$ | 0.9375 | fitted |
| $\omega$ | 1 | fitted |
| $\gamma$ | 0.0261 | fitted |
| $\epsilon$ | 0.0148 | fitted |
| $p$ | 0.0473 | fitted |
| $\phi$ | 0.0265 | fitted |
| $\alpha$ | 0.4927 | fitted |
| $\zeta$ | 5.1462e-04 | fitted |
| $d_1$ | 4.1579e-04 | fitted |
| $d_2$ | 0.0357 | fitted |
| $\nu$ | 0.0377 | fitted |

Figure 4 shows the model projections for COVID-19 daily cases for asymptotic (*I_A_*), self-isolating (*I_AT_*), symptomatic (*I_s_*) and symptomatic isolated (*J*) populations from day one of infection up to day 365. If mass testing were to be stopped after day 87, then more asymptomatic and symptomatic case would remain in the society and undetected. The increase in mass testing after day 87 was associated with a corresponding increase in detected cases and a reduction in undetected cases. Lower peaks of undetected cases were associated increase in mass testing with significant decrease in peak for symptomatic cases.

**Fig. 4.**
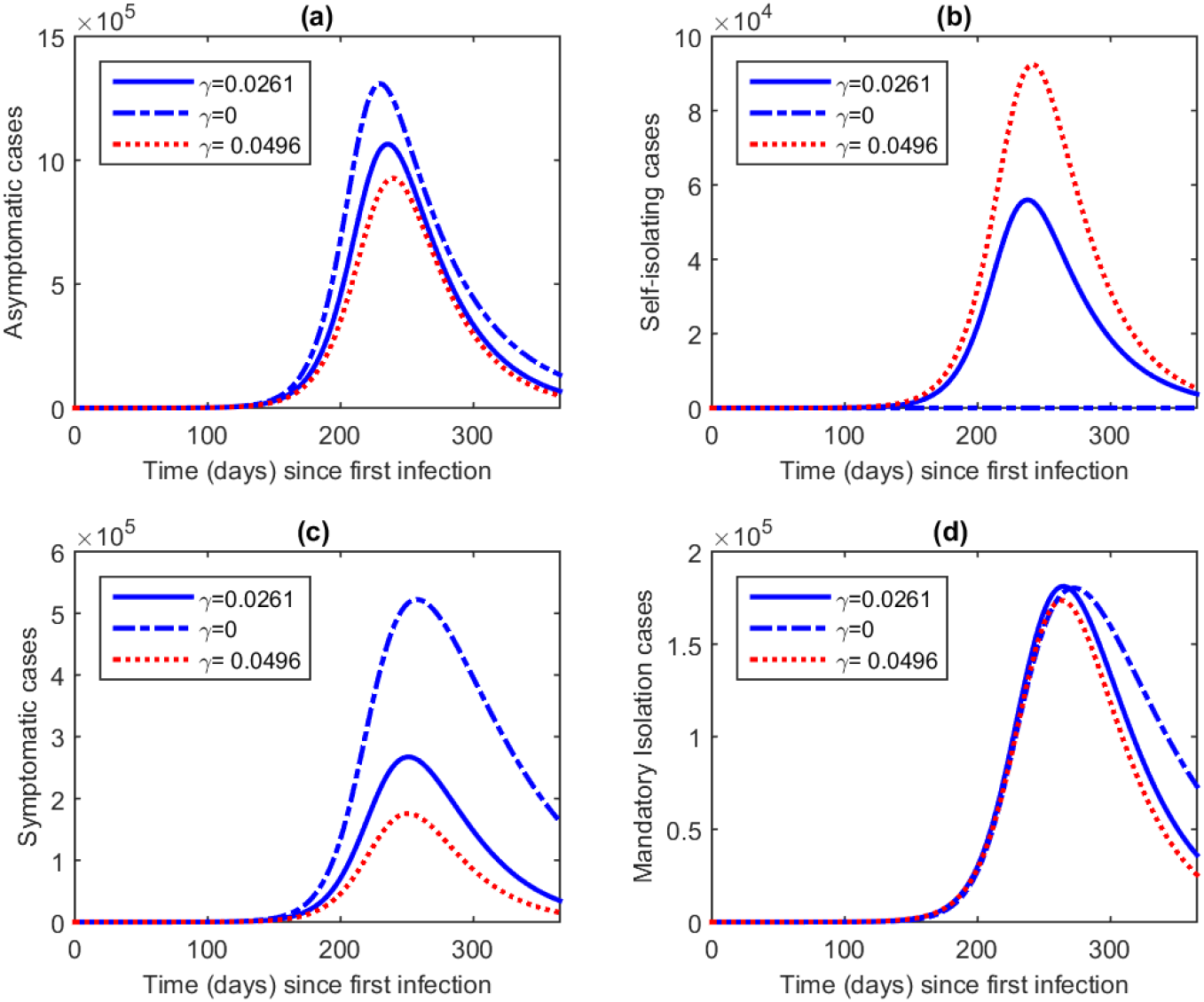
Model projection for COVID-19 daily cases for asymptotic, self-isolating, symptomatic and symptomatic isolated populations from day one of infection up to day 365.

Figure 5 shows model projection for COVID-19 cumulative self-isolating, cumulative admissions from *I_s_*, cumulative admissions from *I_AT_* and overall cumulative admissions for Nigeria from day one of infection up to day 365. We observe that increasing mass testing is associated with increase in cumulative self isolation cases and as well as cumulative admissions of symptomatic cases especially from *I_s_* into monitored isolation centres.

**Fig. 5.**
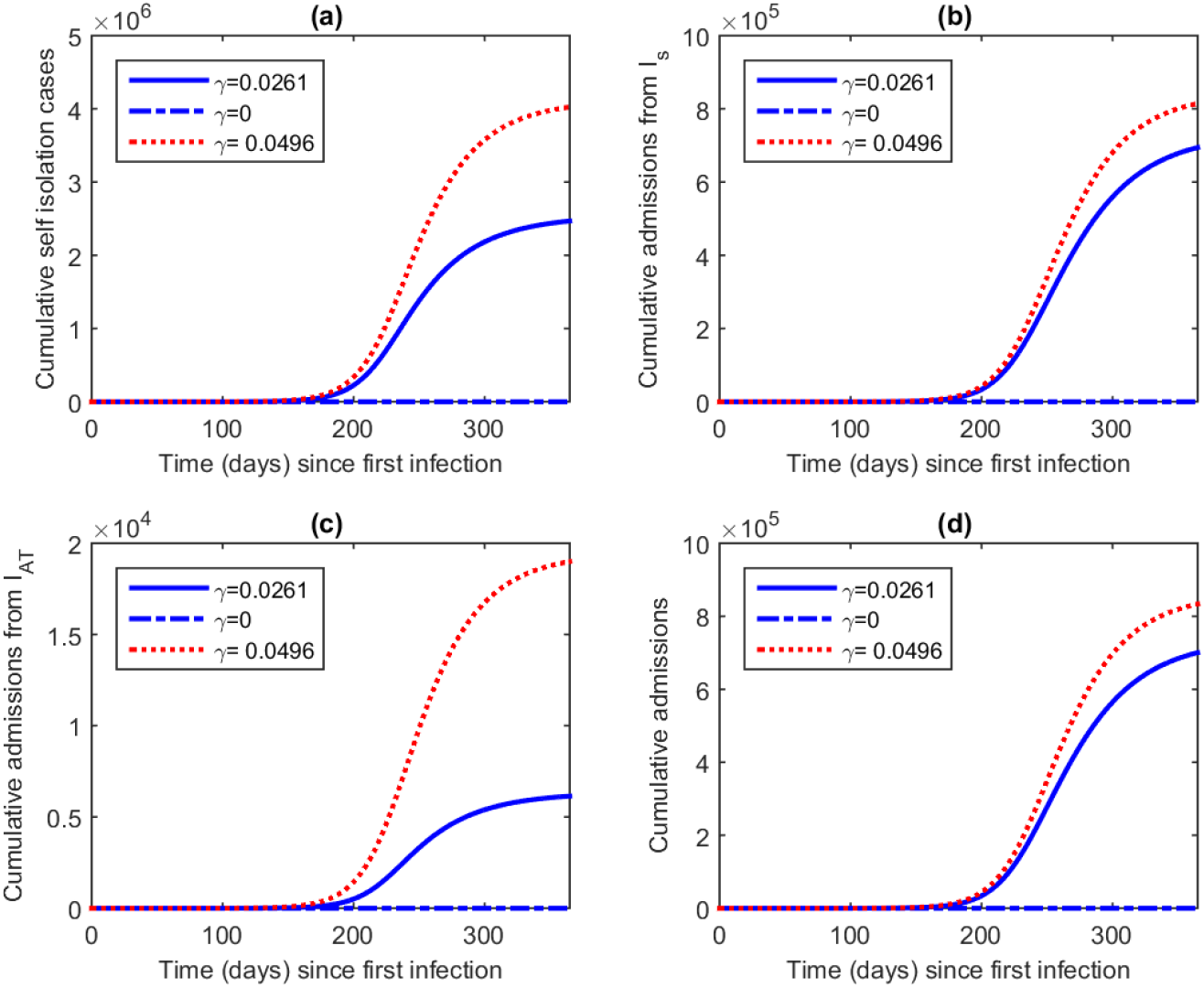
Model projection for COVID-19 cumulative self-isolating, cumulative admissions from *I_s_*, cumulative admissions from *I_AT_* and overall cumulative admissions for Nigeria from day one of infection up to day 365.

Figure 6 shows the model projections for COVID-19 overall cumulative cases, cumulative cases from *I_s_*, cumulative cases from *I_AT_* and cumulative deaths for Nigeria from day one of infection up to day 365. The results show that the cumulative cases will reduce with the increase in mass testing. We also note that more cases will be recorded in the monitored isolation centres. Cumulative deaths will also decrease with increase in mass testing.

The authors declare that they have no conflict of interest.

**Fig. 6.**
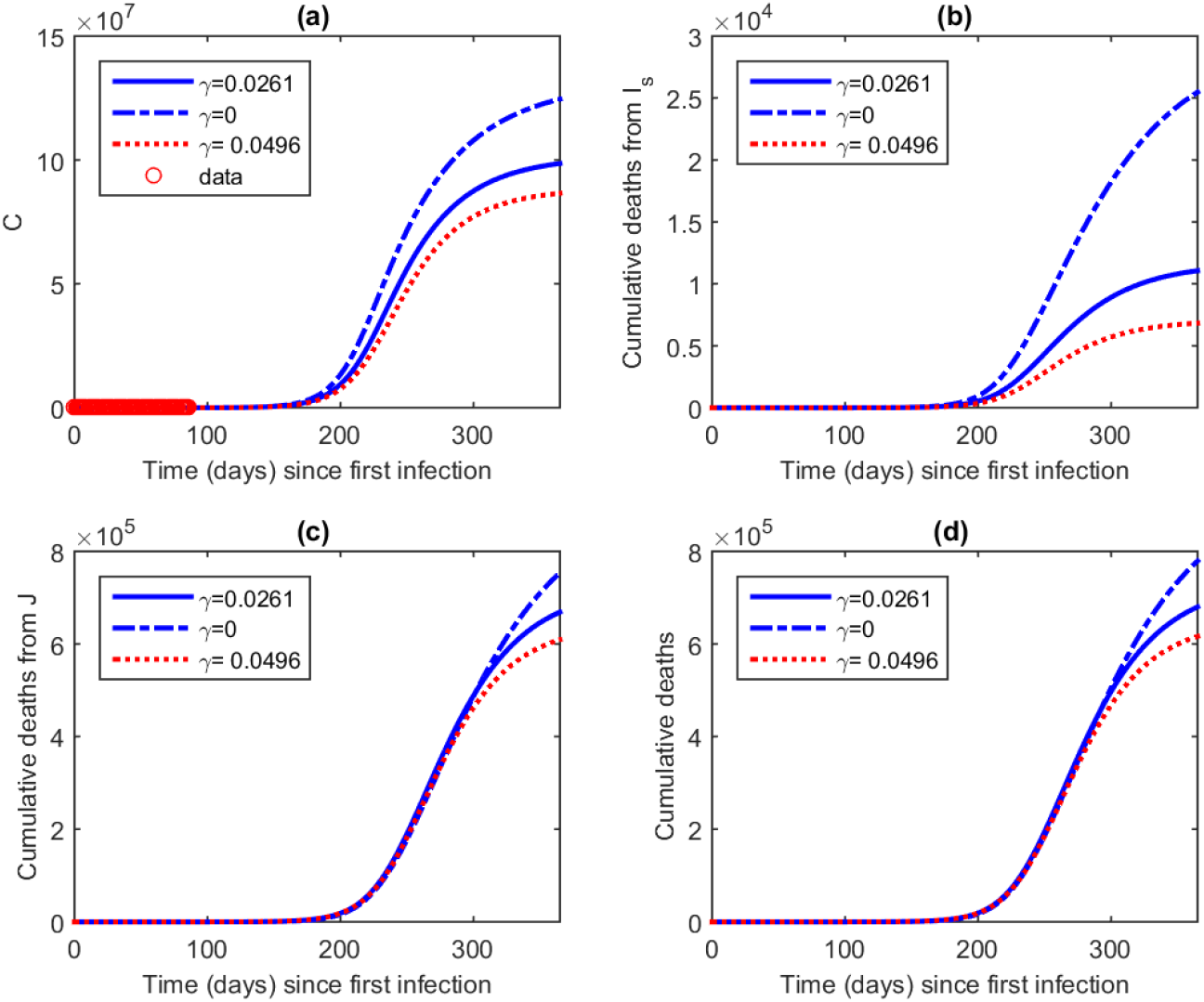
Model projection for COVID-19 overall cumulative cases, cumulative cases from *I_s_*, cumulative cases from *I_AT_* and cumulative deaths for Nigeria from day one of infection up to day 365.

Figure 7 shows the model projections for COVID-19 daily cases for asymptotic, self-isolating, symptomatic and symptomatic isolated populations. The results reveal that better outcomes are achieved when there is increased mass testing and reduced transmission from those that are self-isolating. We also observe that the worst case resulted from stopping mass testing and no transmission from the self-isolating individuals. Figure 8, shows that increasing mass testing and reducing the transmission of self-isolating individuals would initially reduce the cumulative cases but there will be a switch with the baseline scenario. In Figure 9, the worst case which increases cumulative deaths over time is when there is no mass testing and no transmission from self-isolating individuals.

**Fig. 7.**
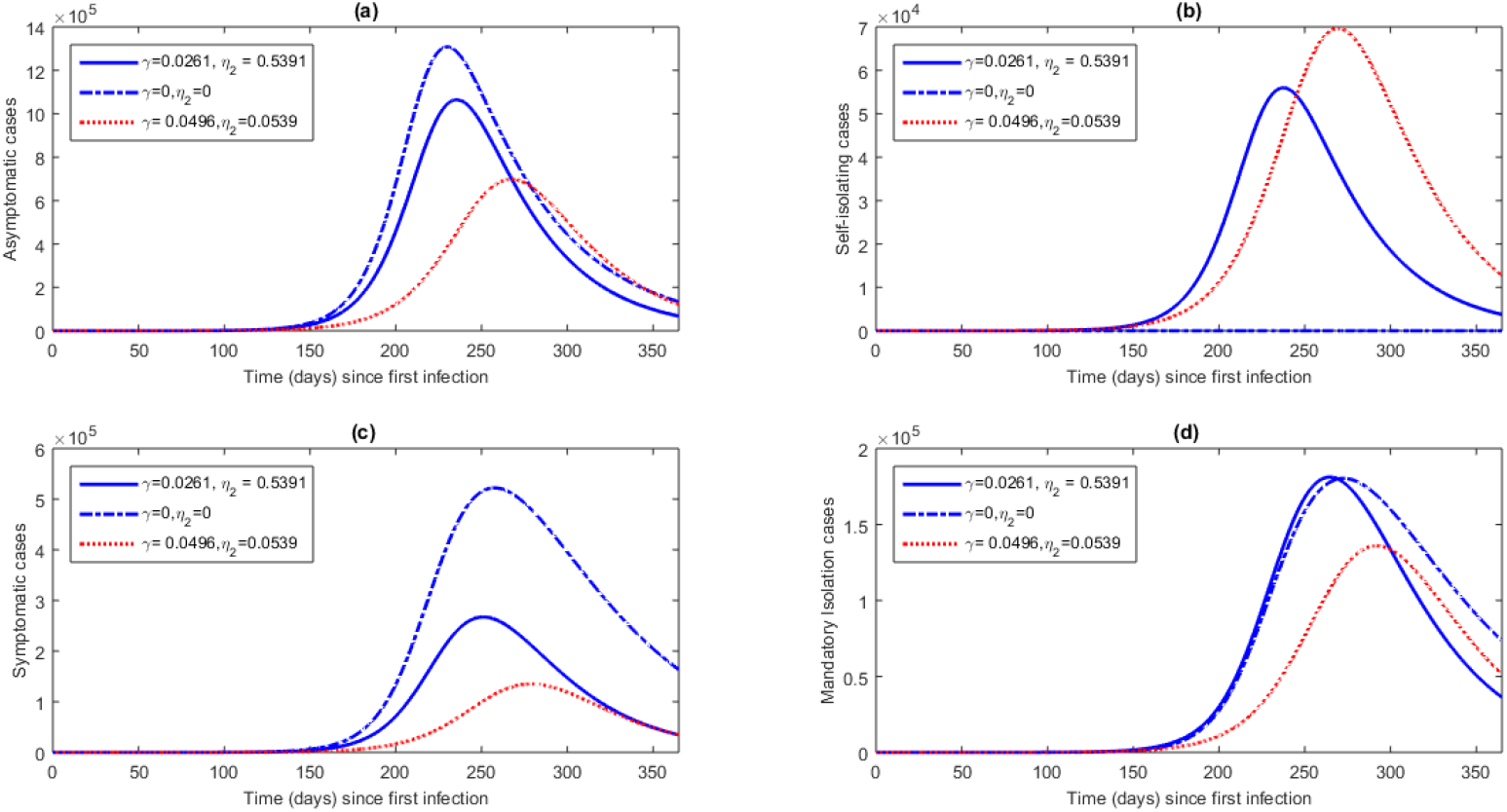
Model projection for COVID-19 daily cases for asymptotic, self-isolating, symptomatic and symptomatic isolated populations from day one of infection up to day 365.

**Fig. 8.**
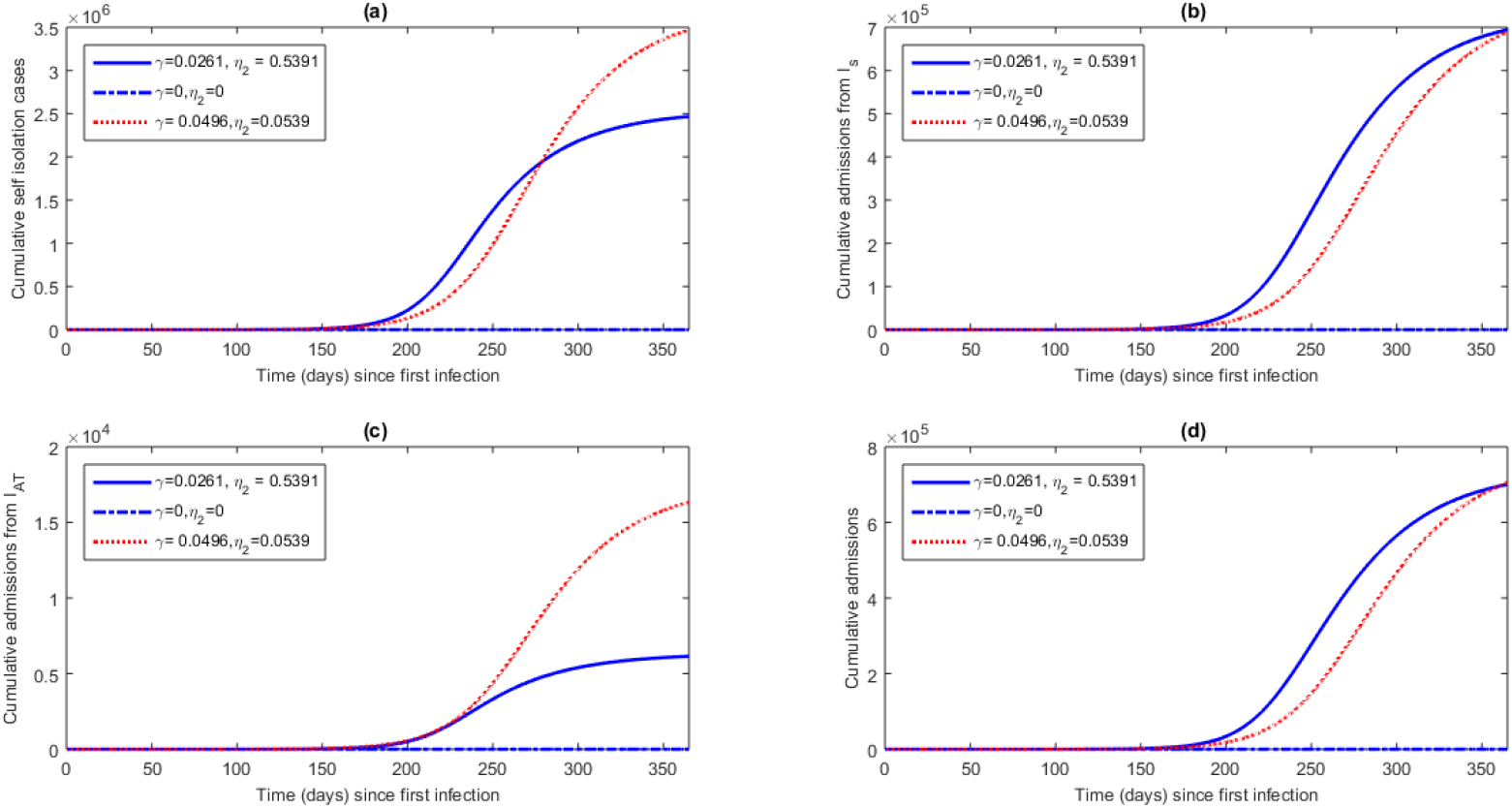
Model projection for COVID-19 cumulative self-isolating, cumulative admissions from *I_s_*, cumulative admissions from *I_AT_* and overall cumulative admissions for Nigeria from day one of infection up to day 365.

**Fig. 9.**
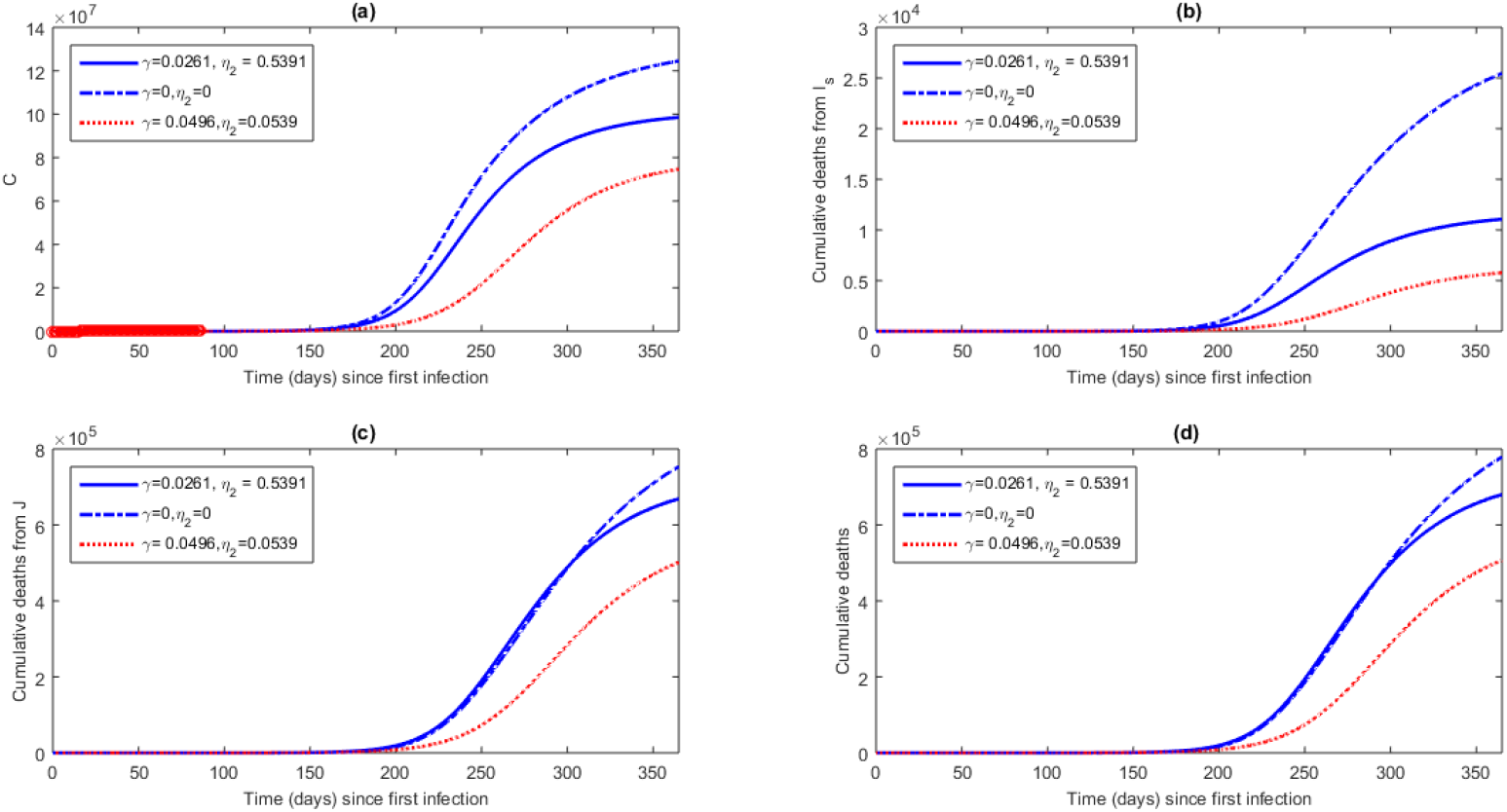
Model projection for COVID-19 overall cumulative cases, cumulative cases from *I_s_*, cumulative cases from *I_AT_* and cumulative deaths for Nigeria from day one of infection up to day 365.

## Conclusion

We presented a mathematical model for COVID-19 for Nigeria using the cumulative cases data to calibrate the model and used the fitted parameters as baseline parameters for model predictions. Mathematical analysis was done and important thresholds for model that can be used to predict the progression or non-progression of the infection in a population. Our results suggest that increase in mass testing is associated with benefits of increasing detected cases, lowering peaks of symptomatic cases, increase in self-isolating cases, decrease in cumulative deaths and decrease in admissions into monitored isolation facilities. If used in combination with monitoring the contact of self-isolating individuals, better benefits are realised when increase in mass testing is done with low transmission rate from the self-isolating individuals but the benefits are short-lived. The worst scenario comes from the case when mass testing is stopped and no transmission from the already self-isolating cases for this is associated with increase in the burden of COVID-19. From our results, we draw the following conclusion: Mass testing is important in exposing asymptomatic cases which should self-isolate and minimize if not have zero transmission. Further, the exposition of these self-isolation cases in turn is associated with lowering of peaks and less clogging of monitored isolation centres as well as the reduction of fatalities. The current mass testing levels in Nigeria have not yet realised these benefits and more efforts should be invested in these strategies to help its health system to manage the worst case scenarios. Lock down measures associated with physical distancing have been shown to delay the peaks and help systems prepare for the worst [23] but they fall short of identifying the potential threat imposed by various infectious groups. The current study has been been able to show similar benefits as the lock down measures but more importantly the specific target groups that could be the silent drivers of infection. With the setup of most under-privileged communities in Nigeria, its impossible to have zero transmission and hence, the strategy of mass testing and isolation should be complemented with other various COVID-19 preventive, management and administrative strategies. Results from this study can easily be adapted to any other country with variables similar to the modelling framework in this study.

## Data Availability

Public data for Nigeria from NCDC (https://covid19.ncdc.gov.ng/)

## Acknowledgements

FC would like to thank his institution for the URC grant and material support that enabled the completion of this project.

